# SARS-CoV-2 circulation in the school setting: A systematic review and meta-analysis

**DOI:** 10.1101/2021.09.03.21263088

**Authors:** Chiara Martinoli, Carlo La Vecchia, Sara Raimondi, Federica Bellerba, Clementina Sasso, Alessandra Basso, Giulio Cammarata, Sara Gandini

**Author notes:** Corresponding author. Department of Experimental Oncology, European Institute of Oncology (IEO), IRCCS, Via Giuseppe Ripamonti 435, 20141 Milan, Italy.

## Abstract

**Background:** The contribution of children to viral spread in schools is still under debate.

We conducted a systematic review and meta-analysis of studies to investigate SARS-CoV-2 transmission in the school setting.

**Methods:** Literature searches from April, 2021 and repeated on May, 15^th^ 2021 yielded a total of 1088 publications: screening, contact tracing and seroprevalence studies.

MOOSE guidelines were followed and data analyzed using random-effects models.

**Results:** From screening studies involving more than 120,000 subjects, we estimated 0.31% (95% Confidence Interval [CI] 0.05-0.81%) SARS-CoV-2 point prevalence in schools. Contact tracing studies, involving a total of 112,622 contacts of children and adults, showed that onward viral transmission was limited (2.54%; 95%CI 0.76-5.31). Young index cases were found to be 74% significantly less likely than adults to favor viral spread (Odds Ratio [OR]=0.26; 95%CI 0.11-0.63) and were less susceptible to infection (OR=0.60; 95% CI 0.25-1.47). Finally, from seroprevalence studies, with a total of 17,879 subjects involved, we estimated that children are 43% significantly less likely than adults to test positive for antibodies (OR=0.57; 95%CI: 0.49-0.68).

In conclusion, testing all subjects in schools, independently of symptoms, students less likely than adults favor viral spread and SARS-CoV-2 circulation in schools was found to be limited.

**KEY POINTS:** *Question:* What is the infectivity and susceptibility of students and staff exposed to SARS-CoV-2 in the school setting?

*Findings:* This systematic review and meta-analysis of all available data shows that SARS-CoV-2 viral spread is limited and child-to-adult transmission in the school setting scarce. Summary estimates indicate that young index cases were 74% significantly less likely than adults to favor viral spread and children are 43% less susceptible than adults.

*Meaning:* Overall, SARS-CoV-2 circulation in schools was limited and could be reasonably controlled with appropriate mitigation measures.

## INTRODUCTION

The global public health crisis due to the severe acute respiratory syndrome coronavirus 2 (SARS-CoV-2) pandemic has brought distinct challenges to the care of children and adolescents globally. School closures have been implemented internationally as a common strategy to control the spread of SARS-CoV-2 during the pandemic, based on the assumption that children may represent important vectors for viral spread. According to UNESCO, 188 countries have imposed countrywide school closures, affecting more than 1.5 billion children and youth, and schooling has been disrupted for an average of 25 weeks worldwide from the beginning of the pandemic until March 2021, due to complete or partial closures (www.unesco.org). The consequences of school closures could be dramatic. It has been estimated that over 100 million children will fall below the minimum proficiency level in reading due to the impact of COVID-19 school closures (www.unesco.org), and children with disabilities and special needs, or living in countries or areas with poor digital connectivity, are especially hard to serve through remote schooling. Beyond providing instruction, school plays a pivotal role in children education, development, and well-being. According to UN reports, over 300 million children rely on school meals for a regular source of daily nutrition, and rising malnutrition is expected among the most vulnerable. Lockdowns and shelter in place measures heightened risk of children witnessing or suffering domestic violence and abuse. Use of online platforms for distance learning has increased risk of exposure to inappropriate content and online predators, and risks to child mental health and well-being are also considerable (www.un.org).

Although data collected from contact-tracing and population studies indicated that children and adolescents are less susceptible to SARS-CoV-2 infection when compared to adults, as shown in a recent meta-analysis,^1^ the contribution of children to viral spread is still under debate. In fact, given the typically mild clinical course of COVID-19 in younger age,^2,3^ symptoms-based testing may have underestimated infection in children, and unrecognized viral circulation may still occur at school, potentially raising community infection rates.

Schools provide a highly regulated environment which is well suited to investigation of potential COVID-19 exposure.^4,5^ Here, we reviewed the current evidence for SARS-CoV-2 transmission in the educational setting. In our analysis, we included studies on point prevalence to assess the potential extent of unrecognized viral positivity in students, serosurveys to estimate the silent circulation of SARS-CoV-2 in schools, and studies on contact tracing to determine the infectivity and susceptibility of students and school staff exposed to the virus.

## METHODS

### Search strategy and selection criteria

We included any original study (article, communication, report), peer-reviewed publications or pre-prints, which reported a quantitative estimation of SARS-CoV-2 transmission in the school setting. We excluded reviews, meta-analyses and modeling studies. Studies performed in special settings were included in the systematic review but excluded from the meta-analysis.

We planned and conducted a systematic literature search and review following MOOSE guidelines regarding meta-analysis of observational studies.

We performed a systematic literature review using validated search strategies from these databases: • PUBMED (http://www.ncbi.nlm.nih.gov/entrez/query.fcgi) • Ovid MEDLINE database • ISI Web of Science: Science Citation Index Expanded (SCI Expanded) and Living Evidence on COVID database (https://zika.ispm.unibe.ch/assets/data/pub/search_beta/) to identify papers on SARS-CoV-2 transmission in the educational setting (Flow-chart Figure 1).

**Figure 1.**
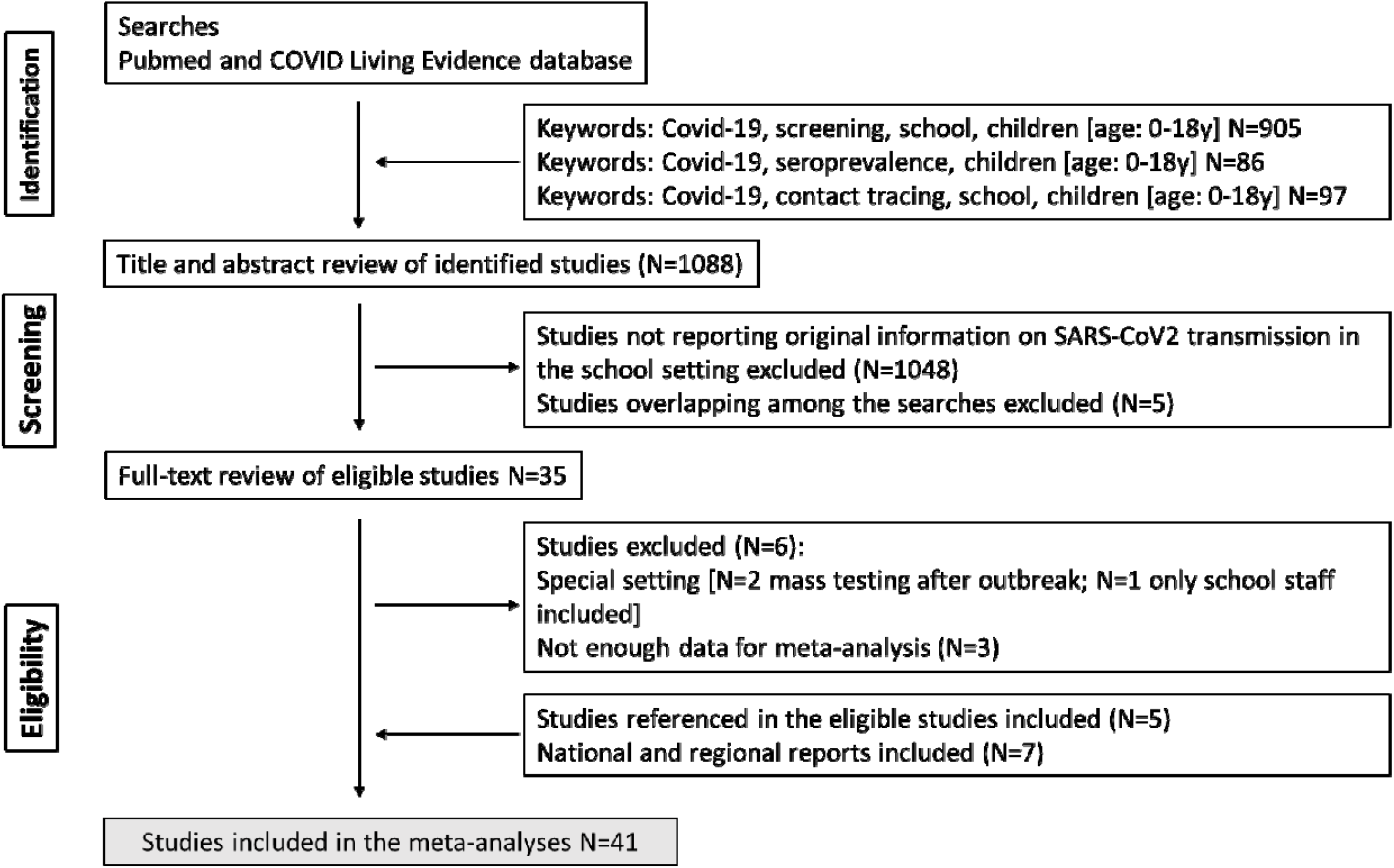
Flow diagram for search.

The search was undertaken during the first week of April, 2021, and repeated on May, 15^th^ 2021. We used the search terms “screening (or point prevalence)”, “serosurvey (or seroprevalence)” and “contact tracing” combined with “school (or education), children”. For PUBMED searches we also limited the search to [age: birth-18y] and added the term “COVID (or COVID-19, or SARS-CoV-2)”. No language restriction was applied. The searches yielded a total of 1088 publications. After screening of titles and abstracts, and removal of duplicates, 35 publications were selected for full-text review. After full-text review, six studies were deemed not eligible for the meta-analysis, and five additional studies were identified after search of cited references in eligible publications. Finally, seven national and regional reports SARS-CoV-2 transmission in the educational setting fulfilling inclusion criteria were identified and included in the meta-analysis.

Data were extracted and cross-checked independently by two investigators. The following information from the published papers was extracted and coded: publication type, country, size of the country, region, study period, study setting, study design, exposure to SARS-CoV-2, test method, test sample, number of subjects tested and testing positive in each category (students or staff), as well as proportions of positive subjects with 95% confidence interval (CI) and confounders adjusted for. In addition, number, age and definition of index cases and their contacts was collected for contact tracing studies.

This study followed the Preferred Reporting Items for Systematic Reviews and Meta-analyses (PRISMA) reporting guideline. Methodological quality of included studies was assessed based on a critical appraisal checklist for prevalence studies (The Joanna Briggs Institute Critical Appraisal tools for use in JBI Systematic Reviews).

### Data analysis

Inclusion criteria: (1) The study contains the minimum information necessary to obtain the percentages of positive subjects, or data to calculate risk estimates, by type of index case, and corresponding 95% CI (i.e., odds ratios or relative risks and a measure of uncertainty: standard errors, variance, confidence intervals or exact P-value of the significance of the estimates); (2) The study is based on independent data to avoid giving double weight to a single study. In case of multiple reports of the same study, we considered the estimates from the most recent publication. Pooled estimates of percentages of positivity were obtained through random-effects models after Freeman-Tukey double arcsine transformation. In case of zero cases Haldane-Anscombe correction was used. All measures of association and the corresponding CI were translated into log relative risk, and corresponding variance, with the formula proposed by Greenland et al.^8^ We used random effects models, taking into account between-study and within-study variability when more than one estimate from a single study was used. Summary Odd Ratio (SOR) was obtained from maximum likelihood estimate: PROC MIXED in SAS, taking into account the model when more estimates were obtained from a single study. Homogeneity of effects across studies was assessed using the Chi-square statistic and quantified by I^2^, which represents the percentage of total variation across studies that is attributable to heterogeneity rather than chance.^9^ We obtained the SOR pooling the study-specific estimates by random effects models. A funnel-plot-based approach was used for assessing publication bias evaluating regression of log(OR) on the sample size, weighted by the inverse of the variance, as suggested by Macaskill et al.^10^

To assess the influence of possible sources of bias, we considered the STROBE (Strengthening the Reporting of Observational Studies in Epidemiology) checklist proposed for observational epidemiologic studies.^11^ According to the STROBE checklist, using meta-regression, we evaluated factors influencing between-study heterogeneity. We also examine changes in results after exclusion of specific studies to evaluate the stability of the pooled estimates. Sensitivity analysis was carried out to evaluate whether results were influenced by single studies.

All the statistical analyses were performed using SAS software (SAS Institute Inc., Cary, NC; version 9.4) and R software, version 4.0.2 (http://www.r-project.org). Two-sided P-value less than 0.05 were considered statistically significant.

## RESULTS

### SARS-CoV-2 positivity rates in educational settings (screening studies)

We identified 21 studies that reported data on SARS-CoV-2 infections in diverse educational settings, from kindergartens and daycares to primary and high schools (eTable 1 in the Supplement),^12–29^ involving more than 120,000 subjects. Fifteen studies were from Europe,^12,13,15– 17,19–24,27,29,30^ five from US,^16,18,25,26,28^ one from Israel.^14^ Overall, studies documented SARS-CoV-2 positivity rates from the beginning of the pandemic in March 2020 until May 2021, including periods of low and high community transmission. Screening campaigns were organized by schools as part of mitigation measures to prevent introduction of SARS-CoV-2 in their premises or by local and national authorities to monitor viral circulation among students, teachers, and non-teaching staff. Testing involved asymptomatic or oligosymptomatic participants, providing an indication of otherwise possibly unrecognized viral spread within educational settings. The vast majority of the studies were judged of high quality (80%, eTable 4). The summary estimate of positivity rate assessed through implementation of the different screening methods was 0.44% (95% CI 0.13-0..92%), with high heterogeneity (*I*^*2*^ = 97%, Figure 2a). The difference in estimates between cross-sectional and cohort studies was significant (p=0.03) with a remarkable lower prevalence among cross-sectional studies (0.31%, 95% CI 0.05-0.81%) compared to cohort studies (1.14%, 95% CI 0.01-4.19%)(Table 1).

**Figure 2a.**
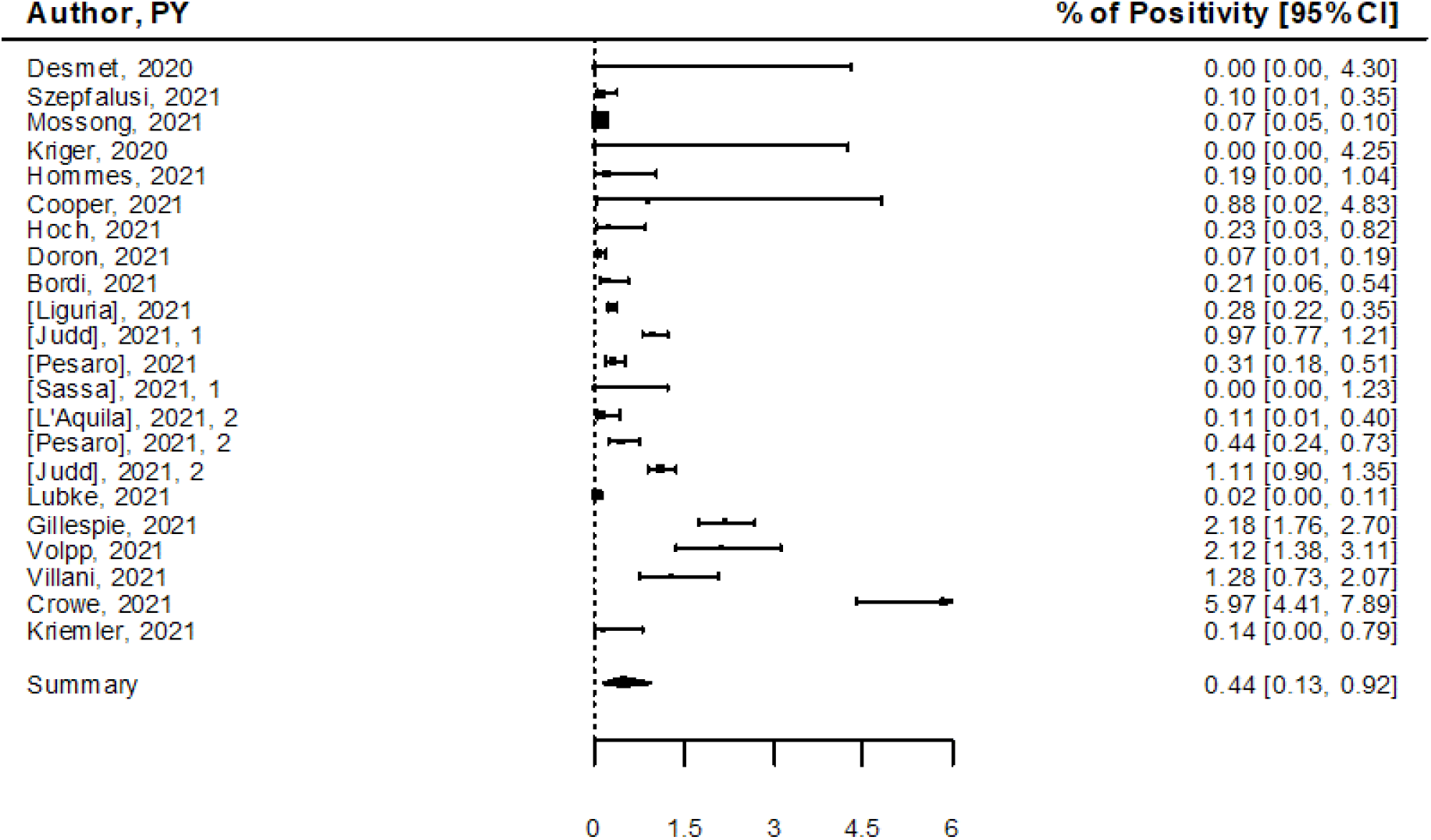
Pooled estimates of testing positive for SARS-CoV-2 (screening studies)

**Figure 2b.**
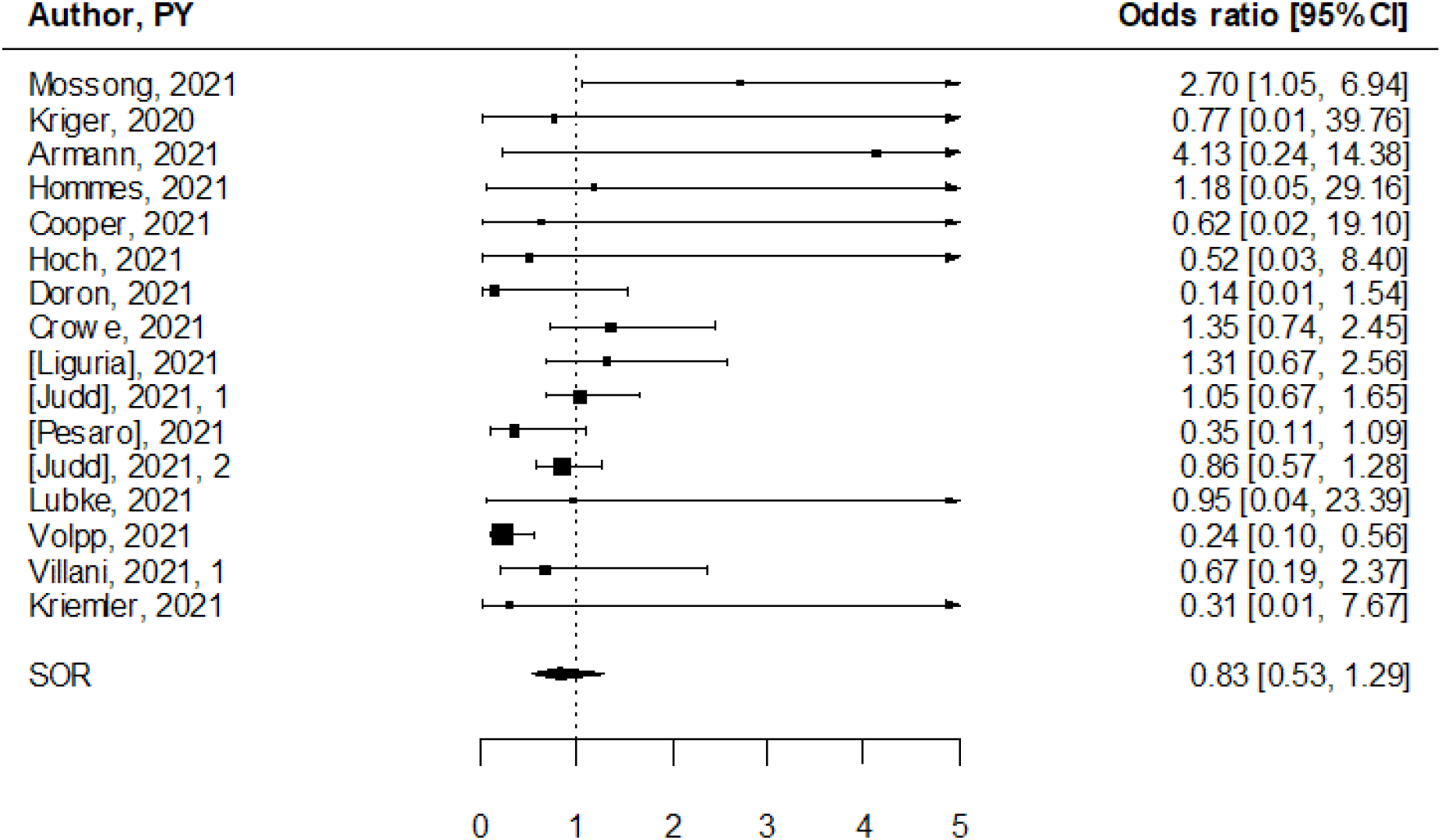
Summary Odds Ratio of testing positive for SARS-CoV-2 for children versus adults linked to educational settings (screening studies)

**Table 1.** Summary estimates.

|  |  | n | Summary | Low<br>95%CI | UP<br>95%CI | I<sup>2</sup> | Study design | P-value |
| --- | --- | --- | --- | --- | --- | --- | --- | --- |
| % Sars-cov2 positivity | Contract tracing* | 15 | 2.54 | 0.76 | 5.31 | 100 |  |  |
|  | Screening | 22 | 0.44 | 0.13 | 0.92 | 97 |  |  |
|  |  | 6 | 1.14 | 0.01 | 4.19 | 98 | Cohorts | 0.03 |
|  |  | 16 | 0.31 | 0.05 | 0.81 | 95 | Cross-sectionals |  |
|  | Serosurvey | 9 | 3.90 | 1.15 | 8.19 | 100 |  |  |
|  |  | 3 | 10.31 | 2.44 | 22.74 | 98 | Cohorts | 0.005 |
|  |  | 6 | 1.49 | 0.07 | 4.69 | 88 | Cross sectionals |  |
| OR for young vs old | Susceptibility in Contract tracing | 6 | 0.60 | 0.25 | 1.47 | 63 |  |  |
|  | Infectivity in Contract tracing | 3 | 0.26 | 0.11 | 0.63 | 44 |  |  |
|  | Screening | 15 | 0.83 | 0.53 | 1.29 | 41 |  |  |
|  |  | 5 | 0.62 | 0.20 | 1.94 | 69 | Cohorts | 0.56 |
|  |  | 10 | 0.98 | 0.74 | 1.32 | 23 | Cross-sectionals |  |
|  | Serosurvey | 6 | 0.57 | 0.46 | 0.68 | 21 |  |  |

\*evaluated considering as denominators contacts not screened subjects; Susceptibility: testing positive young vs adult after a contact case; Infectivity testing positive after a young versus adults contact case; P-value from meta-regression for study design

Fifteen studies reported SARS-CoV-2 point prevalence in a total of 112,131 subjects,^12–23,30–32^ including one study providing data collected during two rounds of testing within the COVID-19 Schools Infection Survey.^23^ A total of 326 coronavirus infections were detected. Overall, estimated positivity rate was 0.31% (95% CI 0 05-0 81%), with high heterogeneity among studies (*I*^*2*^ = 95%, Table 1). Highest infections rates were reported in the two rounds of testing performed in England in November and December 2020, especially among high school students and staff^23^. Excluding reports not published in peer reviewed journals, the summary estimate of positivity rate was slightly greater, 0.40% (95% CI 0.05-1.07%), but still below 1%.

Six cohort studies reported results of multiple testing performed on a total of 12,838 subjects.^24–29^ Multiple testing, especially when intensive schedules over a prolonged period are implemented, may provide indications on the cumulative viral spread within the analyzed educational settings. Overall, estimated positivity rate was 1.14% (95% CI 0.01-4.19%), with high heterogeneity among studies (*I*^*2*^ = 98%, Table 1). Highest number of cases were detected in the study of Crowe and colleagues,^28^ who identified 46 infections in 773 asymptomatic staff and students (5.6%) from three schools engaged in a pilot program with weekly testing over a 5-week period in November 2020 in Omaha, US. Bi-weekly testing led to identification of 25 cases out of 1180 participants (2.1%) over 3-month period in the study held by Volpp and colleagues in New Jersey, US (15-22 samples collected on average from each participant, for a total of 21,449 test performed),^26^ while only 1 case out of 5210 participants (0.02%) was detected in a 4-week study performed in Dusseldorf, Germany (34.068 tested samples, up to 8 tests per subject).^24^ Two rounds of testing one week apart identified one case out of 701 participants in Swiss students and staff participating to Ciao corona study,^29^ while 16 samples tested positive out of the 3431 collected from 1251 students and staff (1.3% positive subjects) of two schools in Rome, Italy, over a 3-month period. Finally, Gillespie and colleagues reported the experience of two US schools that implemented strong mitigation measures for re-opening,^25^ including weekly testing for students and staff. Here, nine rounds of universal testing led to identification of 81 SARS-CoV-2 infected individuals out of the 3720 participants (2.2%), with additional 23 infections identified through contact tracing and 33 self-reported COVID-19 cases.

Nine cross-sectional studies ^13,15–18,20,23,31^ and five cohort studies reported infections detected in children and in adults separately,^24,26–29^ including one reporting estimates for two rounds of testing (eTable 1 in the Supplement).^23^ Children and adults showed comparable SARS-CoV-2 positivity rates in most studies, and the pooled OR estimate was 0.83 (95% CI: 0.53-1.29), with low heterogeneity (*I*^*2*^ = 41%, Figure 2b). We found an indication for publication bias (P=0 03). Similar results were observed in cross-sectional studies (pooled OR = 0 98; 95% CI 0 74-1 32, *I*^*2*^ = 23%), and cohort studies (pooled OR = 0 62; 95% CI 0 20-1 94, *I*^*2*^ = 4%) (Table 1).

### SARS-CoV-2 seroprevalence in educational settings (serosurveys)

We identified 9 studies that reported data on SARS-CoV-2 seroprevalence in educational settings providing in-person activities,^14,15,30,33–38^ with a total of 17,879 subjects involved (eTable 2 in the Supplement). All studies were performed in Europe during 2020. While the above-described point prevalence cross-sectional studies inform on the rates of current infections, cross-sectional serosurveys inform on cumulative exposure to the virus for tested participants. We also identified three cohort studies assessing prevalence of antibodies for SARS-CoV-2 at different time points, with participants tested twice to examine longitudinal changes of seroprevalence,^36–38^ and we included the most recent testing in our metanalysis. The majority of the studies were judged of high quality (70%, eTable 5). The overall seropositive rate was 3.9% (95% CI 1.15-8.19%), with high heterogeneity among studies (*I*^*2*^ = 100%, Figure 3a). The difference in estimates between cross-sectional and cohort studies was statistically significant (p=0.005), with estimates obtained from cohort studies indicating a 10% of positivity (10.3%; 95% CI 2.43-22.7%) and a lower prevalence of 1.5% from cross-sectional studies (1.49%, 95% CI 0.07-4.69%) (Table 1).

**Figure 3a.**
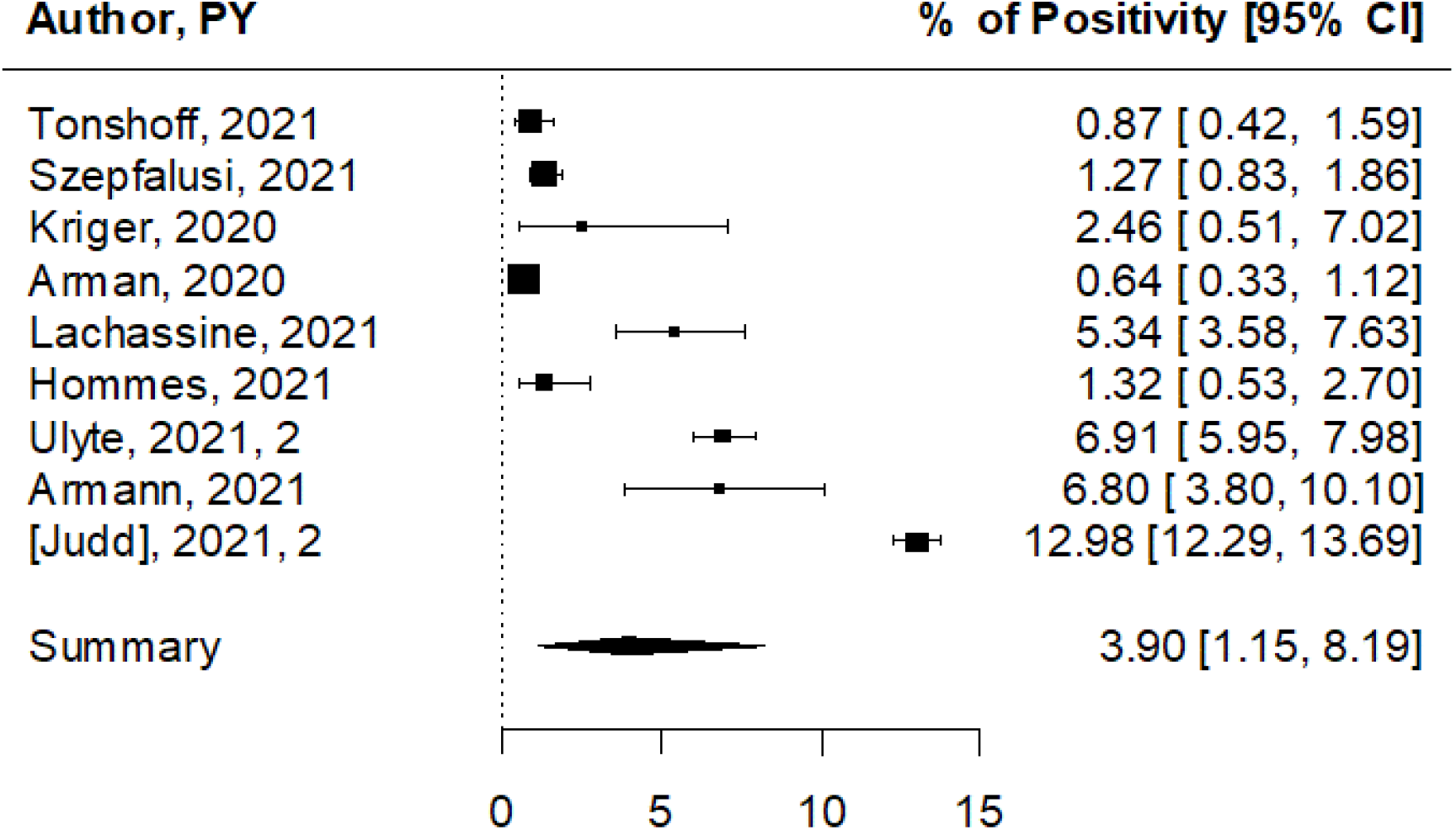
Pooled estimates of testing positive for antibodies for SARS-CoV-2 (Serosurveys)

**Figure 3b.**
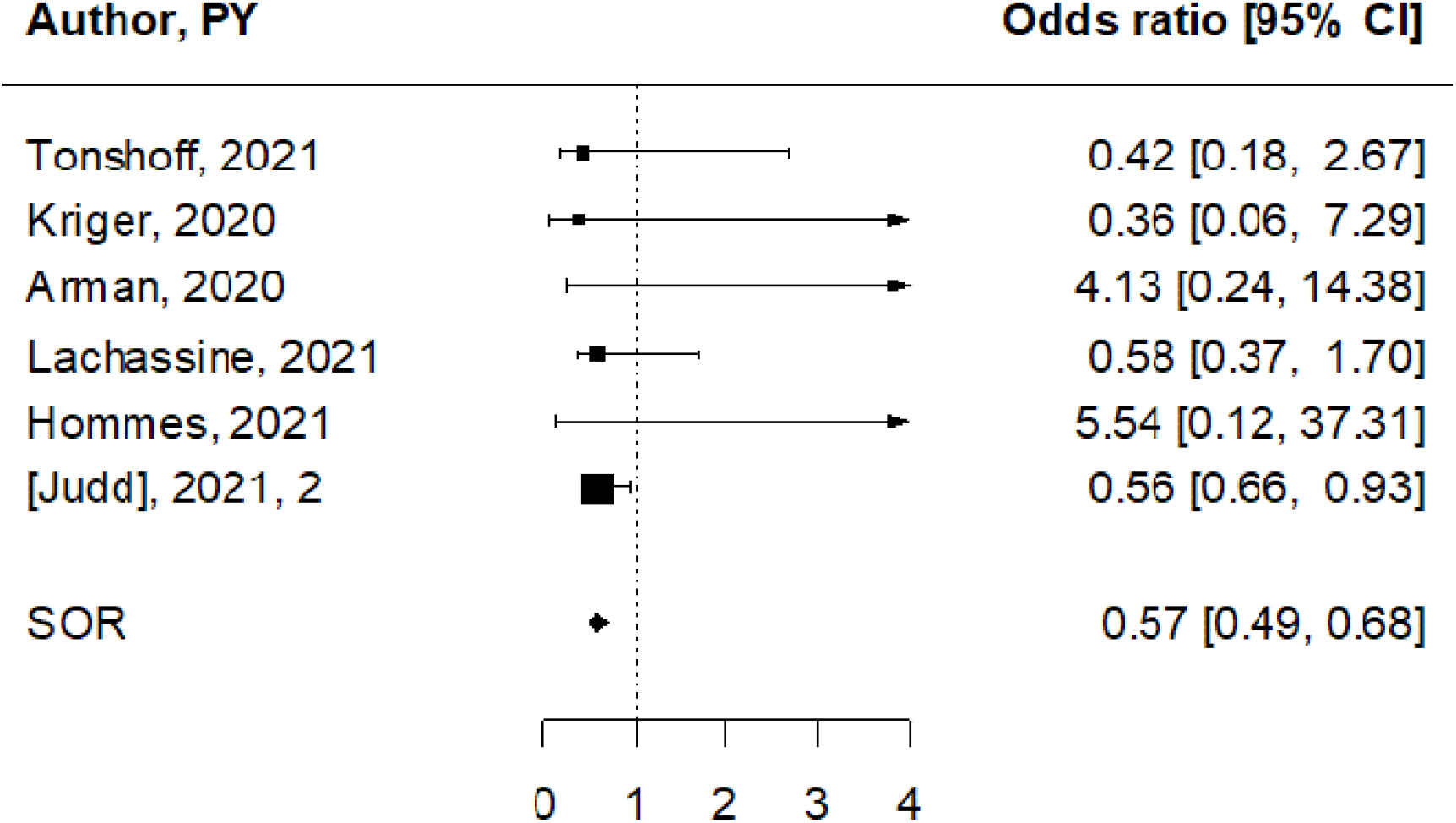
Pooled estimates of odds of testing positive for antibodies for SARS-CoV-2 for children versus adults (serosurveys)

Six studies were performed during the first semester of 2020 and assessed antibodies during or shortly after the first wave of the pandemic,^14,15,30,34,35^ including three studies focused on attendant to schools and daycares that remained exceptionally open during national lockdowns.^14,33,35^ Although the seroprevalence differed among the three studies, authors reported comparable seroprevalence in groups of children not attending school (0.5% versus 1%, and 1.4% versus 2.7%, respectively, for children attending in-person, or staying at home),^14,33^ and adults who did not have occupational contact with children or COVID-19-positive patients (7.7% for daycare staff and 5.5% for the comparator adult group,^35^ suggesting that exceptional schooling did not boost SARS-CoV-2 spread in the analyzed settings). In line with these observations, low seroprevalence reported in three additional cross-sectional studies adds up to the indication that schools did not develop into silent hotspots for viral transmission during the first wave of the pandemic,^15,30,34^ likely due to the successful implementation of extensive preventive measures. In this regard, it is noteworthy mentioning two studies reporting seroprevalence in students from a small city in north France and from a large community school in Santiago, Chile.^39,40^ In both cases, school-based COVID-19 outbreaks occurred at the very onset of the pandemic and in the absence of preventive measures, leading to 38.1% and 43.4% seropositive pupils and teachers, respectively, in one French high school,^39^ and 9 9% and 16 6% seropositive students and staff in the Chilean study.^40^

In the three cohort studies,^36–38^ increased seroprevalence was recorded during the second study visit, which took place after the summer break,^36^ or in December 2020.^37,38^ In their study, Ulyte and colleagues reported that SARS-CoV-2 seroprevalence raised from 3% in the summer to 4.5% in late autumn in school children in the canton of Zurich, Switzerland.^36^ Interestingly, among the children who participated to both testing rounds, 28/70 (40%) who were initially seropositive became seronegative, while seroconversion (previously seronegative participants who became seropositive) was 5% (109/2153). The estimated rate of ever-positive children was therefore 7.8%.^36^ Serial blood sampling was also implemented in a secondary school in Dresden, Germany.^37^ Here, antibody positivity rates increased from 1.7% to 6.8% during the 6-week study period, and all the participants who tested positive at the initial visit (5/5) remained positive at the second visit. Lastly, data collected within the COVID-19 Schools Infection Survey showed high initial antibody positivity rates, consistent with the study designed to oversample schools in areas of England where coronavirus infection was highest at the start of the academic year (September 2020), and a not significant increase in pupils (7.7% to 9% and 11% to 13.5% for primary and secondary school, respectively) and staff (12.5% to 15%) testing positive for SARS-CoV-2 antibodies between November and December 2020. In this study, 7.3% (20/276) of staff who initially tested positive, had no detectable antibodies in the second round.

Separate seroprevalence estimates for children and adults were available for six studies (eTable 2 in the Supplement).^14,15,33,37,38,41^ School-aged children had lower antibody positivity rates when compared with adults (parents or school staff) in 4/6 studies (Figure 3b). The pooled OR for children was 0.57 (95% CI 0.49-0.68), significantly lower than adults, with low heterogeneity. among studies (*I*^*2*^ = 21%). No indication for publication bias was found (P=0 42).

### Onward transmission of SARS-CoV-2 in educational settings (contact tracing)

Studies analyzing onward transmission of SARS-CoV-2 offered the possibility to test more specifically the infectivity and susceptibility to infection of children linked to educational settings. We identified 15 studies that reported data on transmission of SARS-CoV-2 in schools with available information on the number of contacts of index cases (eTable 3 in the Supplement).^14,42– 53^Six studies were from Europe,^43,45,47,50,51,54^ five from USA,^41,48,49,52,53^ two from Israel,^14,46^ one each from South Korea and Australia.^42,44^ A total of 112,622 contacts of children and adults who were physically present at school while positive for SARS-CoV-2, were identified. Molecular testing for SARS-CoV-2 was normally offered to all contacts exposed to SARS-CoV-2, except for one study^48^. In this study, only symptomatic contacts were tested, so asymptomatic secondary cases were not captured. The majority of the studies were judged of high quality (60%, eTable 6).

When considering any-age index cases, and their contacts of any age, the pooled secondary attack rate (SAR) was 2 54% (95% CI 0 76-5 31%), with high heterogeneity among studies (*I*^*2*^ = 100%, Figure 4a). The highest attack rate (13.5%) was recorded within a large outbreak in a high school in Jerusalem, Israel linked to two student index cases and likely promoted by inadequate preventive measures (crowded classes, exemption from facemasks and continuous air-conditioning due to an extreme heatwave).^46^ High attack rates were recorded also in two US-based studies, both reporting transmission with index cases of any age. Doyle and colleagues investigated COVID-19 in primary and secondary schools in Florida during the first semester of school reopening.^49^ Of the 63,654 of COVID-19 cases registered between August and December 2020 in school-aged children, 60% were not school-related, and <1% of registered students were identified as having school-related COVID-19. Contact tracing investigations identified 86,832 persons who had close school setting contact with these cases; among these, 37,548 received testing and 10,092 received a positive SARS-CoV-2 test result, leading to 11.6% secondary attack rate. Prospective investigation of SARS-CoV-2 transmission was also performed in a Georgia school district during a period of peak community COVID-19 incidence.^52^ Tracing of 86 index cases identified 1,005 contacts, of whom 644 were tested and 59 received a positive SARS-CoV-2 test result (SAR=5.9%). Highest SAR were identified in the setting of indoor sports and staff interactions. On the other hand, extremely low attack rates (<1%) were found in five studies reporting from five nations and two continents and describing transmission of SARS-CoV-2 in school setting from the early onset of the pandemic (one pediatric case among the first reported cases in France who visited three schools and one ski class while infected, generated 172 contacts and one secondary case) until November 2020.^43^ Similar results were found in the study conducted in Italy during the second wave, September to November 2020^55^. Secondary infections at school were <1% and secondary infections among teachers were rare, occurring more frequently when the index case was a teacher than a student (37% vs. 10%, P = 0.007).

**Figure 4a.**
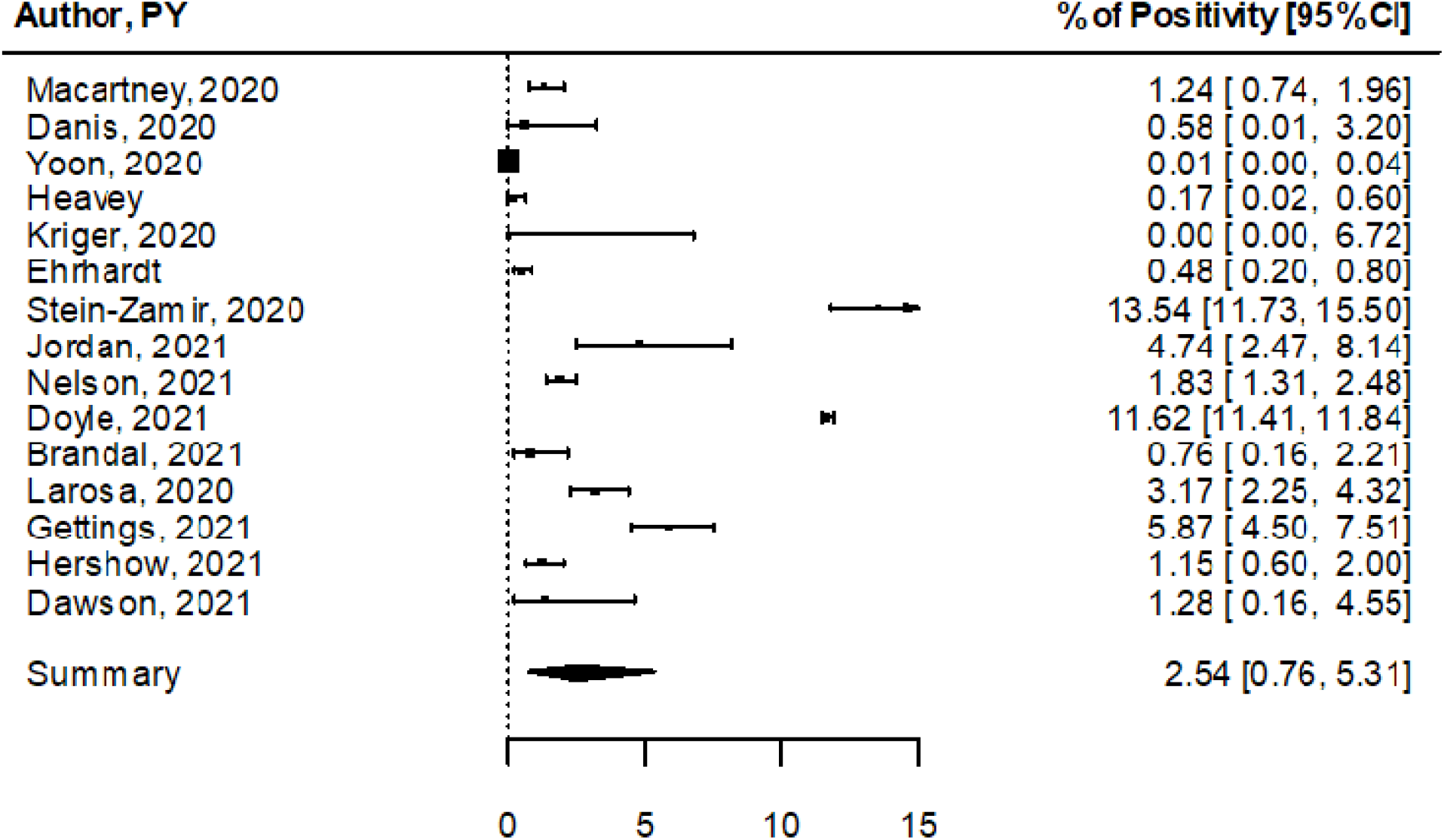
Pooled estimates of testing positive for SARS-CoV-2 after a contact with a case in the school setting (contact tracing studies)

**Figure 4b.**
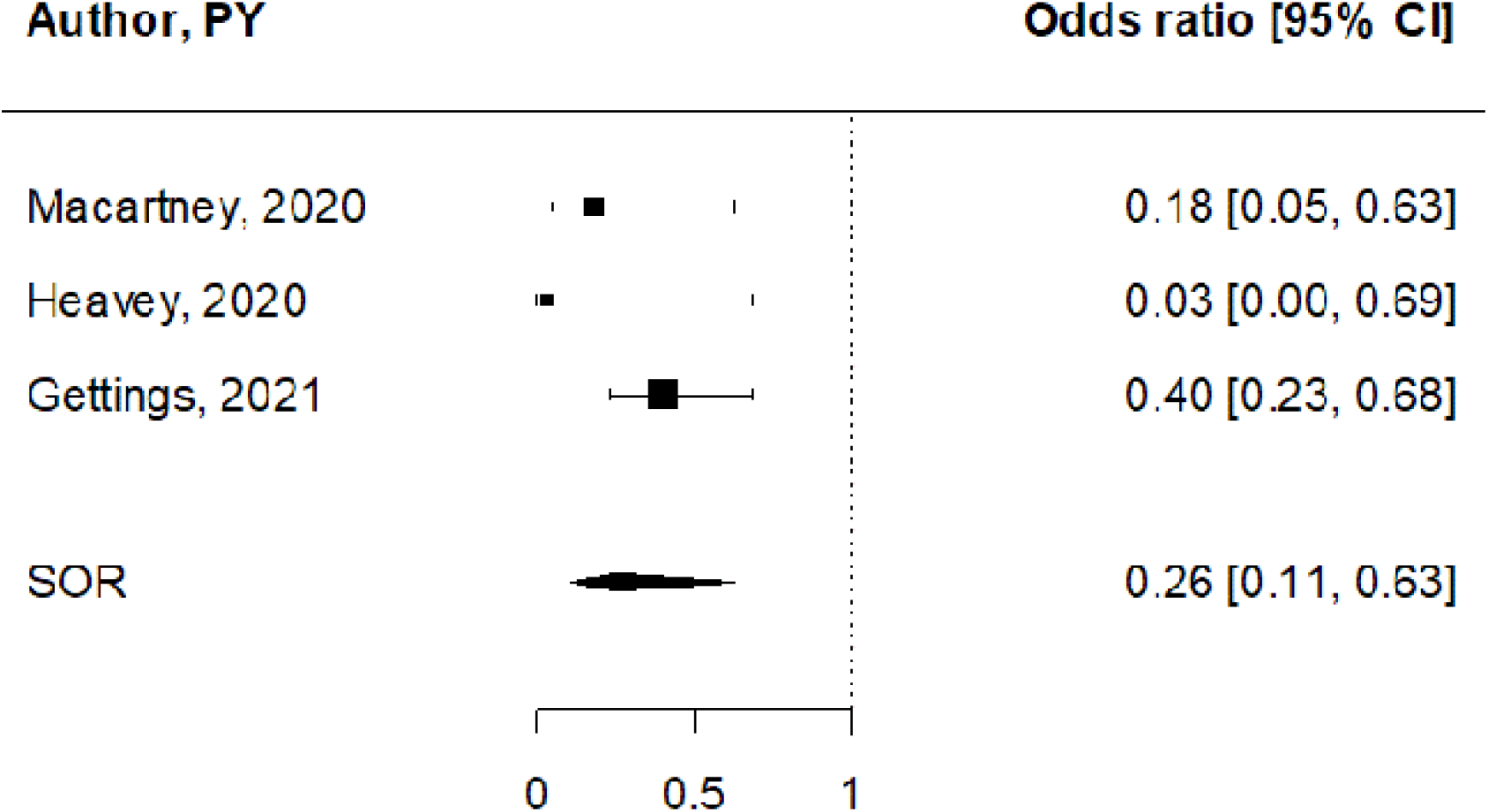
Summary OR of testing positive for SARS-CoV-2 after a contact with a young versus an adult case in the school setting (contact tracing studies)

**Figure 4c.**
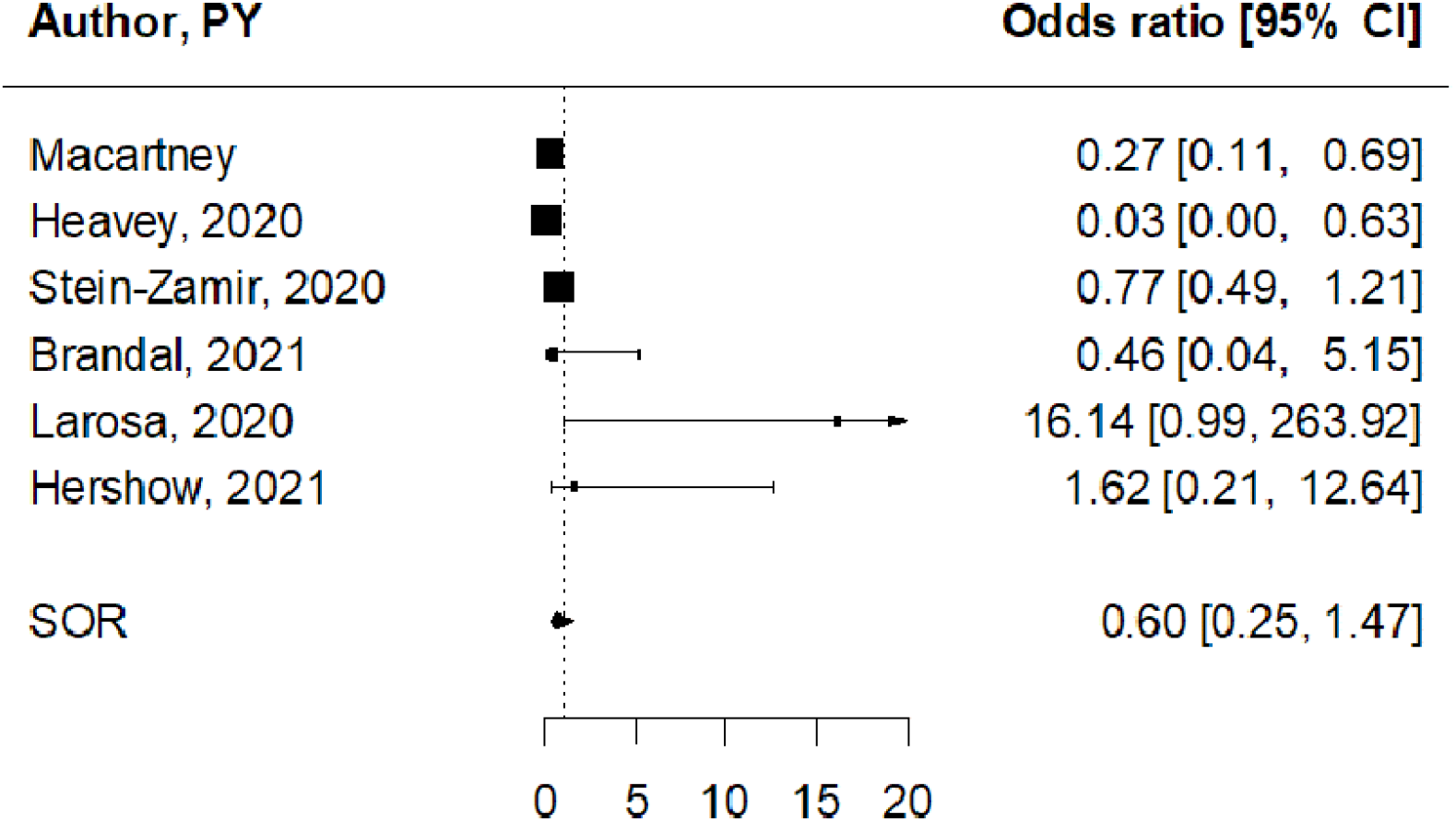
Summary OR of testing positive for SARS-CoV-2 after a young versus adults contact case.

Three studies reported on viral transmission from young (age <18y) or adult index cases (eTable 3 in the Supplement).^42,52,54^ In all studies, the proportion of contacts infected by young index cases was lower compared to adult index cases (Figure 4b). The pooled OR was 0.26 (95% CI 0.11-0.63), indicating a significant 3-fold reduced infectivity for children compared with adults. No indication for publication bias was found (P=0.59).

Six studies reported estimates of viral transmission to young versus adult contacts (eTable 3 in the Supplement).^42,46,50,51,53,54^ The pooled OR was 0 60 (95% CI 0 25-1 47), indicating a not statistically significant reduced susceptibility to SARS-CoV-2 infection for children compared to adults (Figure 4c). No indication for publication bias was found (P=0.65).

## Discussion

The findings of this systematic review and meta-analysis suggest that schools did not develop into hotspots for SARS-CoV-2 transmission. Although infection can and does occur in schools, more than 1-year experience with in-person schooling worldwide indicates limited viral spread when mitigation measures are adopted, with low SARS-CoV-2 circulation and scarce child-to-adult transmission in the school setting.

Reviewing the experience of different schools showed that health behavioral policies realized before the vaccination, seem to mitigate the risk of viral spread in educational settings. In particular contact tracing is very useful to promptly isolate infected staff and students. We found five times greater frequency of positive tests with contact tracing compared to screening and these results suggests that testing all subjects in schools, independently of symptoms, probably not help in reducing clusters. Finally, it would be pivotal to prioritize teachers and educational support staff in COVID-19 vaccine rollout, especially older people and subjects with chronic diseases, to protect teachers ‘and students’ health and safety.

Engagement of all stakeholders (school staff members, students, and their parents or legal guardians) into implementation of school-based policies, as well as individual adherence to shared recommendations are key for minimizing COVID-19 transmission chain.

## Data Availability

Data available on request from the authors

## FIGURES

**Figure 1. Flow diagram for search**

**Figure 2 Pooled estimates for SARS-CoV-2 screening studies**

a) Pooled estimate of testing positive for SARS-CoV-2.

b) Summary Odds Ratio of testing positive for SARS-CoV-2 for children versus adults linked to educational settings.

**Figure 3 Pooled estimates for SARS-CoV-2 seroprevalence studies**

a) Pooled estimate of testing positive for antibodies for SARS-CoV-2.

b) Pooled estimate of odds of testing positive for antibodies for SARS-CoV-2 for children versus adults.

**Figure 4 Pooled estimates for contact tracing studies**

a) Pooled estimate of testing positive for SARS-CoV-2 after a contact with a case in the school setting.

b) Summary OR of testing positive for SARS-CoV-2 after a contact with a young versus an adult case in the school setting.

c) Summary OR of testing positive for SARS-CoV-2 in young versus adults after a contact with a case.

## Data sharing

The study dataset can be requested from Sara Gandini upon request.

## Acknowledgements

The European Institute of Oncology, Milan, Italy is partially supported by the Italian Ministry of Health with Ricerca Corrente and 5×1,000 funds.

Federica Bellerba is a Ph.D. student within the European School of Molecular Medicine (SEMM).

The funder of the study had no role in study design, data collection, data analysis, data interpretation, or writing of the report.

