## Supplementary tables for "SARS-CoV-2 circulation in the school setting: A systematic review and meta-analysis"

**eTable 1: Studies on screening for SARS-CoV-2 infections**

| **First author**  **[report]** | **Country** | **Study design** | **Start obs** | **Stop obs** | **Test method** | **Test sample** | **Subjects tested** | **N tested** | **N pos** |
| --- | --- | --- | --- | --- | --- | --- | --- | --- | --- |
| Desmet^12^ | Belgium | Cross-sectional | 02/03/2020 | 12/03/2020 | RT-PCR | Swab | Children  (Daycare 6-30 mo) | 84 | 0 |
| Szépfalusi^30^ | Austria | Cross-sectional | 01/05/2020 | 30/07/2020 | RT-PCR | Swab | Students (5-21y) | 2064 | 2 |
| Mossong^13^ | Luxembourg | Cross-sectional | 04/05/2020 | 25/07/2020 | RT-PCR | Swab | Students and staff  Students  Staff | 48380  33723  14657 | 36  31  5 |
| Kriger^14^ | Israel | Cross-sectional | 07/05/2020 | 17/05/2020 | RT-PCR | Swab | Children and parents  Daycare (8±2.5 y)  Parents | 85  48  37 | 0  0  0 |
| Hommes^15^ | Germany | Cross-sectional | 11/06/2020 | 19/06/2020 | RT-PCR | Swab | Students and staff  Primary school students  High school students  Staff | 532  193  192  150 | 1  0  1  0 |
| Cooper^16^ | USA | Cross-sectional | 01/09/2020 | 01/11/2020 | RT-PCR | Swab | Students and staff  K12 Students  Staff | 113  86  27 | 1  1  0 |
| Hoch^17^ | Germany | Cross-sectional | 07/09/2020 | 30/11/2020 | RT-PCR | Saliva and swab | Students and staff  Students (3-12y)  Staff | 875  574  301 | 2  1  1 |
| Doron^18^ | USA | Cross-sectional | 01/10/2020 | 08/10/2020 | RT-PCR | Saliva | Students and staff  K12 students  Staff | 4601  3596  1005 | 3  1  2 |
| Bordi^19^ | Italy | Cross-sectional | 06/10/2020 | 02/11/2020 | RT-PCR | Saliva | Students 2-15y | 1905 | 4 |
| [Liguria]^20^ | Italy | Cross-sectional | 09/10/2020 | 14/05/2021 | RDT | Swab | Students and staff  Primary school students  High school students  Staff | 25961  15191  6237  4533 | 72  44  18  10 |
| [Judd]^23^ | England | Cross-sectional (Round 1) | 03/11/2020 | 19/11/2020 | RT-PCR | Swab | Students and staff  Primary school students  High school students  Staff | 7923  1936  2420  3567 | . |
| [Pesaro]^31^ | Italy | Cross-sectional | 28/01/2021 | 03/02/2021 | RDT | . | Students and staff  High school students  Staff | 5104  4568  536 | 16  12  4 |
| [L'Aquila]^22^ | Italy | Cross-sectional | 19/02/2021 | 20/02/2021 | RDT | Swab | Students and staff | 1800 | 2 |
| [Pesaro]^21^ | Italy | Cross-sectional | 04/03/2021 | 12/03/2021 | RDT | Swab | Students and staff | 3200 | 14 |
| [Sassa]^32^ | Italy | Cross-sectional | 18/05/2021 | 18/05/2021 | RDT | Swab | Students and staff | 299 | 0 |
| [Judd]^38^ | England | Cross-sectional (Round 2) | 03/11/2020 | 19/11/2020 | RT-PCR | Swab | Students and staff  Primary school students  High school students  Staff | 8650  1898  2988  3764 | . |
| Lübke^24^ | Germany | Cohort | 10/06/2020 | 07/07/2020 | RT-PCR | Saliva | Children and staff  Daycare (0-7y)  Staff | 5210  3955  1255 | 1  1  0 |
| Gillespie*^25^ | USA | Cohort | 5/8/2020 | 20/12/2020 | RT-PCR | Swab  or saliva | Students and staff  K12 Schools | 3720 | 81* |
| Volpp^26^ | USA | Cohort | 20/08/2020 | 27/11/2020 | RT-PCR | Swab | Students and staff  Students  Staff | 1180  775  405 | 25  8  17 |
| Villani^27^ | Italy | Cohort | 21/09/2020 | 04/12/2020 | RT-PCR | Saliva | Students and staff  Students  Staff | 1251  1083  168 | 16  13  3 |
| Crowe^28^ | USA | Cohort | 09/11/2020 | 11/12/2020 | RT-PCR | Saliva | Students and staff  K12 Students  Staff | 770  315  455 | 46  22  24 |
| Kriemler^29^ | Switzerland | Cohort | 01/12/2020 | 11/12/2020 | RT-PCR | Swab | Students and staff  Students (9-14y)  Staff | 707  641  66 | 1  1  0 |

*Additional school cases: 23 contact tracing, 33 self-reported

**eTable 2. Studies on serosurveys for antibodies to SARS-CoV-2**

| **First author**  **[report]** | **Country** | **Study design** | **Start obs** | **Stop obs** | **Test method** | **Test sample** | **Subjects tested** | **N tested** | **N pos** |
| --- | --- | --- | --- | --- | --- | --- | --- | --- | --- |
| Tönshoff^33^ | Germany | Cross-sectional | 22/04/2020 | 15/05/2020 | Quantitative | Serum | Students and parents  Students  Parents | 1156  580  576 | 10  3  7 |
| Szépfalusi^30^ | Austria | Cross-sectional | 01/05/2020 | 30/07/2020 | Quantitative | Serum | Students (5-21y) | 2042 | 26 |
| Kriger^14^ | Israel | Cross-sectional | 07/05/2020 | 17/05/2020 | Quantitative | Serum | Children and parents  Daycare (8±2.5 y)  Parents | 122  70  52 | 3  1  2 |
| Armann^34^ | Germany | Cross-sectional | 25/05/2020 | 30/06/2020 | Quantitative | Serum | Students and staff  High school students  Staff | 1865  1358  507 | 12  11  1 |
| Lachassinne^35^ | France | Cross-sectional | 04/06/2020 | 03/07/2020 | Qualitative | Finger prick blood | Children and staff  Daycare (0-5 y)  Parents | 524  327  197 | 28  14  14 |
| Hommes^15^ | Germany | Cross-sectional | 11/06/2020 | 19/06/2020 | Quantitative | Finger prick blood | Students and staff  Students  Staff | 531  382  149 | 7  7  0 |
| Ulyte^36^ | Switzerland | Cohort_T1 | 16/07/2020 | 09/07/2020 | Quantitative | Serum | Students | 2496 | 74 |
| Ulyte^36^ | Switzerland | Cohort_T2 | 26/10/2020 | 18/11/2020 | Quantitative | Serum | Students | 2503 | 173 |
| Armann^37^ | Germany | Cohort_T1 | Nov2020 | Nov2020 | Quantitative | Serum | Students and staff  (High school) | 302 | 5 |
| Armann^37^ | Germany | Cohort_T2 | Dec2020 | Dec2020 | Quantitative | Serum | Students and staff  (High school) | 273 | 16 |
| [Judd]^38^ | England | Cohort_T1 | 03/11/2020 | 19/11/2020 | Quantitative | Serum | Students and staff  Primary school students  High school students  Staff | 7719  1996  2449  3274 | . |
| [Judd]^38^ | England | Cohort_T2 | 03/11/2020 | 19/11/2020 | Quantitative | Serum | Students and staff  Primary school students  High school students  Staff | 8899  2152  3280  3467 | . |

**eTable 3. Studies on contact tracing**

|  |  |  |  |  |  |  | **All contacts** | | **Contacts <18y** | | **Adult contacts** | |
| --- | --- | --- | --- | --- | --- | --- | --- | --- | --- | --- | --- | --- |
| **First author** | **Country** | **Start obs** | **Stop obs** | **Contacts** | **Age index** | **N index** | **N** | **N pos** | **N** | **N pos** | **N** | **N pos** |
| Macartney^42^ | Australia | 25/01/2020 | 01/05/2020 | Community | Any  <18y  Adult | 27  12  15 | 1448  752  696 | 18  3  15 | .  649  536 | .  2  8 | .  103  160 | .  1  7 |
| Danis^43^ | France | 31/01/2020 | 14/02/2020 | In-school | <18y | 1 | 172 | 1 | . | . | . | . |
| Yoon^44^ | SK | 18/02/2020 | 31/07/2020 | In-school | <18y | 44 | 13100 | 1 | . | . | . | . |
| Heavey^54^ | Ireland | 01/03/2020 | 13/03/2020 | Community | Any  <18y  Adult | 6  3  3 | 1165  999  166 | 2  0  2 | .  905  106 | .  0  0 | .  94  60 | .  0  2 |
| Kriger^14^ | Israel | 07/05/2020 | 17/05/2020 | In-school | Adult | 1 | 53 | 0 | . | . | 53 | 0 |
| Ehrhardt^45^ | Germany | 25/05/2020 | 05/08/2020 | In-school | <18y | 137 | 2300 | 11 | . | . | . | . |
| Stein-Zamir^46^ | Israel | 28/05/2020 | 30/05/2020 | In-school | <18y | . | 1315 | 178 | 1164 | 153 | 152 | 25 |
| Jordan^47^ | Spain | 29/06/2020 | 31/07/2020 | In-school | Any | 39 | 253 | 12 | . | . | . | . |
| Nelson^48^ | USA | 01/08/2020 | 30/11/2020 | In-school | <18y | 257 | 2189 | 40 | . | . | 2189 | 40 |
| Doyle^49^ | USA | 10/08/2020 | 21/12/2020 | Community | Any | . | 86832 | 10092 | . | . | . | . |
| Brandal^50^ | Norway | 28/08/2020 | 11/11/2020 | In-school | <18y | 13 | 393 | 3 | 319 | 2 | 74 | 1 |
| Larosa^51^ | Italy | 01/09/2020 | 15/10/2020 | In-school | Any | 48 | 1200 | 38 | 204 | 0 | 996 | 38 |
| Gettings^52^ | USA | 01/12/2020 | 22/01/2021 | In-school | Any  <18y  Adult | 86  53  33 | 1005  689  421 | 59  24  35 | . | . | . | . |
| Hershow^53^ | USA | 03/12/2020 | 31/01/2021 | In-school | Any | 51 | 1041 | 12 | 908 | 11 | 133 | 1 |
| Dawson^41^ | USA | 07/12/2020 | 18/12/2020 | In-school | Any | 37 | 156 | 2 | . | . | . | . |

**eTable 4: Studies on screening for SARS-CoV-2 infections: quality evaluation**

| **First author** | 1. Was the sample frame appropriate to address the target population? | 2. Were study participants sampled in an appropriate way? | 3. Was the sample size adequate? | 4. Were the study subjects and the setting described in detail? | 5. Was the data analysis conducted with sufficient coverage of the identified sample? | 6. Were valid methods used for the identification of the condition? | 7. Was the condition measured in a standard, reliable way for all participants? | 8. Was there appropriate statistical analysis? | 9. Was the response rate adequate, and if not, was the low response rate managed appropriately? | Quality |
| --- | --- | --- | --- | --- | --- | --- | --- | --- | --- | --- |
| Desmet^12^ | 0 | 1 | 0 | 1 | 0 | 1 | 1 | 0 | 1 | Low |
| Szépfalusi^30^ | 1 | 1 | 1 | 1 | 0 | 1 | 1 | 0 | 1 | High |
| Mossong^13^ | 1 | 1 | 1 | 1 | 1 | 1 | 1 | 0 | 1 | High |
| Kriger^14^ | 0 | 1 | 0 | 1 | 0 | 1 | 1 | 0 | 1 | Low |
| Hommes^15^ | 1 | 1 | 0 | 1 | 0 | 1 | 1 | 0 | 1 | medium |
| Cooper^16^ | 0 | 1 | 0 | 1 | 0 | 1 | 1 | 0 | 1 | Low |
| Hoch^17^ | 1 | 1 | 1 | 1 | 1 | 1 | 1 | 0 | 1 | High |
| Doron^18^ | 1 | 1 | 1 | 1 | 1 | 1 | 1 | 0 | 1 | High |
| Bordi^19^ | 1 | 1 | 1 | 1 | 0 | 1 | 1 | 0 | 1 | High |
| [Liguria]^20^ | 1 | 1 | 1 | 1 | 1 | 0 | 1 | 0 | 1 | High |
| [Judd]^23^ | 1 | 1 | 1 | 1 | 1 | 1 | 1 | 0 | 1 | High |
| [Pesaro]^31^ | 1 | 1 | 1 | 1 | 1 | 0 | 1 | 0 | 1 | High |
| [L'Aquila]^22^ | 1 | 1 | 1 | 1 | 1 | 0 | 1 | 0 | 1 | High |
| [Pesaro]^21^ | 1 | 1 | 1 | 1 | 1 | 0 | 1 | 0 | 1 | High |
| [Sassa]^32^ | 0 | 1 | 0 | 1 | 0 | 0 | 1 | 0 | 1 | Low |
| [Judd]^38^ | 1 | 1 | 1 | 1 | 1 | 1 | 1 | 0 | 1 | High |
| Lübke^24^ | 1 | 1 | 1 | 1 | 1 | 1 | 1 | 1 | 1 | High |
| Gillespie*^25^ | 1 | 1 | 1 | 1 | 1 | 1 | 1 | 1 | 1 | High |
| Volpp^26^ | 1 | 1 | 1 | 1 | 1 | 1 | 1 | 1 | 1 | High |
| Villani^27^ | 1 | 1 | 1 | 1 | 1 | 1 | 1 | 1 | 1 | High |
| Crowe^28^ | 1 | 1 | 1 | 1 | 1 | 1 | 1 | 1 | 1 | High |
| Kriemler^29^ | 1 | 1 | 1 | 1 | 1 | 1 | 1 | 1 | 1 | High |

**eTable 5: Studies on serosurveys for antibodies to SARS-CoV-2: quality evaluation**

| **First author** | 1. Was the sample frame appropriate to address the target population? | 2. Were study participants sampled in an appropriate way? | 3. Was the sample size adequate? | 4. Were the study subjects and the setting described in detail? | 5. Was the data analysis conducted with sufficient coverage of the identified sample? | 6. Were valid methods used for the identification of the condition? | 7. Was the condition measured in a standard, reliable way for all participants? | 8. Was there appropriate statistical analysis? | 9. Was the response rate adequate, and if not, was the low response rate managed appropriately? | Quality |
| --- | --- | --- | --- | --- | --- | --- | --- | --- | --- | --- |
| Tönshoff^33^ | 1 | 1 | 1 | 1 | 1 | 1 | 1 | 0 | 1 | High |
| Szépfalusi^30^ | 0 | 0 | 1 | 1 | 0 | 1 | 0 | 0 | 1 | Low |
| Kriger^14^ | 0 | 0 | 0 | 1 | 0 | 1 | 0 | 0 | 0 | Low |
| Armann^34^ | 1 | 1 | 1 | 1 | 1 | 1 | 1 | 0 | 1 | High |
| Lachassinne^35^ | 1 | 1 | 1 | 1 | 1 | 0 | 1 | 0 | 1 | High |
| Hommes^15^ | 1 | 1 | 1 | 1 | 1 | 0 | 1 | 0 | 1 | High |
| Ulyte^36^ | 0 | 0 | 1 | 1 | 0 | 1 | 0 | 1 | 1 | Medium |
| Ulyte^36^ | 0 | 0 | 1 | 1 | 0 | 1 | 0 | 1 | 1 | Medium |
| Armann^37^ | 1 | 1 | 0 | 1 | 1 | 1 | 1 | 1 | 0 | High |
| Armann^37^ | 1 | 1 | 0 | 1 | 1 | 1 | 1 | 1 | 0 | High |
| [Judd]^38^ | 1 | 1 | 1 | 1 | 1 | 1 | 1 | 1 | 1 | High |
| [Judd]^38^ | 1 | 1 | 1 | 1 | 1 | 1 | 1 | 1 | 1 | High |

**eTable 6: Studies on contact tracing: quality evaluation**

| **First author** | 1. Was the sample frame appropriate to address the target population? | 2. Were study participants sampled in an appropriate way? | 3. Was the sample size adequate? | 4. Were the study subjects and the setting described in detail? | 5. Was the data analysis conducted with sufficient coverage of the identified sample? | 6. Were valid methods used for the identification of the condition? | 7. Was the condition measured in a standard, reliable way for all participants? | 8. Was there appropriate statistical analysis? | 9. Was the response rate adequate, and if not, was the low response rate managed appropriately? | Quality |
| --- | --- | --- | --- | --- | --- | --- | --- | --- | --- | --- |
| Macartney^42^ | 0 | 1 | 1 | 1 | 1 | 0 | 1 | 1 | 1 | High |
| Danis^43^ | 1 | 0 | 0 | 1 | 0 | 1 | 1 | 0 | 1 | Medium |
| Yoon^44^ | 1 | 1 | 1 | 1 | 0 | 1 | 1 | 0 | 1 | High |
| Heavey^54^ | 0 | 1 | 1 | 1 | 1 | 0 | 1 | 1 | 1 | High |
| Kriger^14^ | 1 | 0 | 0 | 1 | 0 | 1 | 1 | 0 | 1 | Medium |
| Ehrhardt^45^ | 1 | 1 | 1 | 1 | 0 | 1 | 1 | 0 | 1 | High |
| Stein-Zamir^46^ | 1 | 0 | 1 | 1 | 0 | 1 | 1 | 1 | 1 | High |
| Jordan^47^ | 1 | 1 | 0 | 1 | 1 | 1 | 0 | 0 | 1 | Medium |
| Nelson^48^ | 1 | 1 | 1 | 1 | 0 | 1 | 1 | 0 | 1 | High |
| Doyle^49^ | 0 | 0 | 1 | 1 | 1 | 0 | 0 | 0 | 1 | Low |
| Brandal^50^ | 1 | 0 | 0 |  | 0 | 1 | 1 | 1 | 1 | Medium |
| Larosa^51^ | 1 | 1 | 1 |  | 1 | 1 | 0 | 1 | 1 | High |
| Gettings^52^ | 1 | 1 | 1 | 1 | 1 | 1 | 1 | 0 | 1 | High |
| Hershow^53^ | 1 | 1 | 1 | 1 | 1 | 1 | 0 | 1 | 1 | High |
| Dawson^41^ | 1 | 1 | 0 | 1 | 1 | 1 | 0 | 0 | 1 | Medium |
